# Integrative analyses of neuroimaging, clinical, and multi-omics data identified meaningful Alzheimer’s disease progression subtypes

**DOI:** 10.1101/2022.11.01.22281820

**Authors:** Qiyuan An, Mengliang Zhang, Xinyue Hu, Chang Su, Fei Wang, Yingying Zhu

## Abstract

Alzheimer’s Disease (AD) is a common neurodegenerative disorder with diverse clinical manifestations. To better understand differences between diverse manifestations, we investigate from the difference in MRI ROI volumes among subtypes to their related different gene pathways. In this study, we identify three subtypes of AD development with fast, moderate, and slow progression rates from the MRI ROI volume biomarkers in the ADNI cohort. We further find differentially expressed proteins in CSF fluids among these AD patients and related significantly different genetic pathways. A total of 159 patients’ data are included for analysis. We identify three distinct subtypes from the hierarchical clustering on MRI ROI volume biomarkers. Subtype 1, characterized as fast progression, consists of 34 patients (21.9%). Subtype 2, as moderate progression, comprises 77 patients (49.6%). Subtype 3, as slow progression, consists of 44 patients (28.3%). These subtypes show significantly differences in cognitive test scores and corresponding genetic pathways.

## 1 Introduction

Alzheimer’s Disease (AD) is a complex and prevalent neurodegenerative disease, whose etiology remains unclear to date. It’s heterogeneous in distinct clinical manifestations, progression trajectories, volume biomarker readouts, and genetic pathways. Due to its irreversible and age-related characters and lack of any cure, early diagnosis and intervention of AD deterioration play an significant role in designing clinical treatment strategies. One major challenge for AD prediction and customized clinical treatment is to identify the phenotypic heterogeneity within the AD population ^i^.

Distinguishing subtypes in AD may lead to insights of different etiological mechanisms, better the customized treatments, and interfere the development of brain atrophy and dementia. The motivation of this study is to investigate into the progression patterns of brain region volume biomarkers among different participants in ADNI cohort, to reveal the underlying atrophy development patterns among different subtypes. We cluster the participant features and identify three subtypes with distinct AD progression patterns in the ADNI cohort: subtype 1, fast symptom progression; subtype 2, moderate symptom progression; and subtype 3, slow symptom progression.

Some previous attempts about neurodegenerative disease subtyping can be categorized into the following categories: using neuroimaging biomarkes’ readouts to identify distinct patterns of atrophy regions ^ii iii^, using cognitive test scores ^iv v^, and using proteomics data^vivii^. However, none of these works take advantage of all these features and derive the underlying significant genetic pathways.

To this end, we propose to identify subtypes of AD via the neuroimaging biomarkers, find differentially expressed proteins from Cerebrospinal Fluid (CSF) proteomics data, and perform pathway enrichment analysis on omics data to derive significant gene pathways. We hope this study can provide insights to discover individually customized treatment for AD patients.

## 2 Experiments

### 2.1 Data Preprocessing

We adopt the ADNI TADPOLE challenge dataset^viii^ for our experiment. The TADPOLE standard dataset includes the following biomarkers:

Main cognitive tests – neuropsychological tests administered by a clinical expert:

- CDR Sum of Boxes
- ADAS 11
- ADAS 13
- MMSE
- RAVLT
- MOCA
- Ecog

MRI ROIs (Freesurfer) - measures of brain structural integrity:

- Volumes
- Cortical thicknesses
- Surface areas

We only adopt the MRI ROI volumes as inputs for subtyping, while using cognitive tests to validate the subtyping results. The number of MRI ROI volumes is 270. The TADPOLE dataset includes 1610 participants, which consists of 342 AD, 417 CN, and 872 MCI. Out of 342 AD participants, we filter out participants with less than 12 months longitudinal data and result in 313 participants. After that, we build each participant’s speed feature defined as 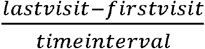, representing the progression rates of different ROI volumes. Then we filter the speed feature matrix in both the participants and the ROI volume dimensions if the missing rate is greater than 0.7. Filtering speed features results in 239 columns ROI volumes and 159 participants. In the end, we perform minmax normalization on the speed feature matrix.

### 2.2 Matrix Decomposition

We decompose the speed matrix via Truncated SVD with rank of 3 to derive participants’ features. The choice of Truncated SVD is for its native suitability for sparse matrix as our speed feature matrix has many missing values. The actual implementation of decomposition is via the sklearn library^ix^.

### 2.3 Clustering

Then we cluster decomposed participants’ features via the hierarchical clustering to avoid the biased/wrong prior hypothesis of number of subtypes. Our clustering results in 3 phenotypes (number of participants) that correspond to fast progression (34), moderate progression (44), and mild progression (77). We validate the subtyping results on cognitive test scores shown in Fig 1.

**Figure 1.**
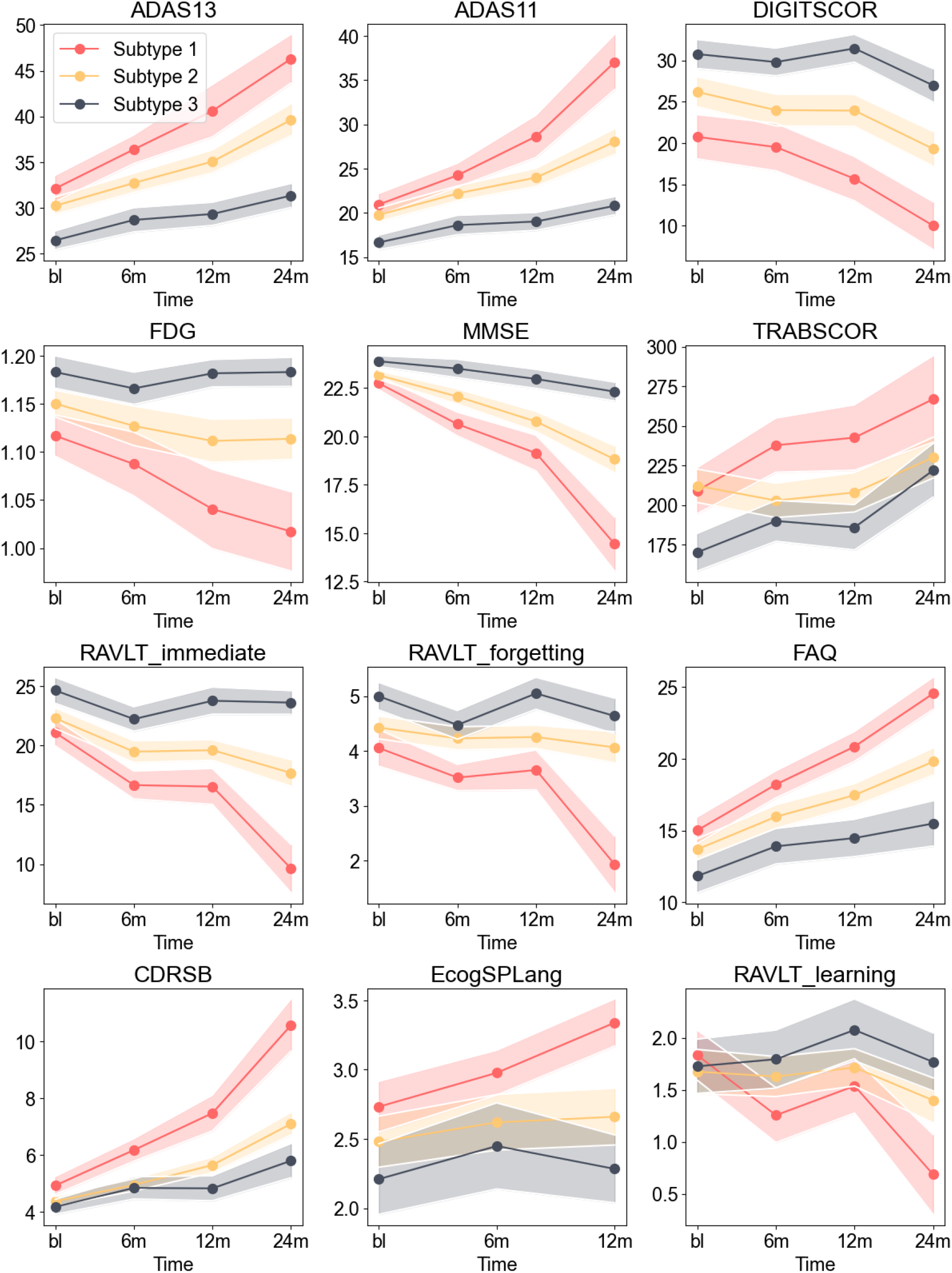
Cognitive test scores in 3 subtypes. Red: subtype 1, yellow: subtype 2, gray: subtype 3.

Each strip in Fig 1 represents the longitudinal trend on one cognitive test of one subtype.

The solid line in the center is the mean of the cognitive test, while the light color between the solid line is the standard error. The clear separability of cognitive scores justifies the legitimacy of our method on different progression trends.

### 2.4 Significance test on ROI volumes

We perform the Kruskal-Wallis test among ROI volumes under different subtypes and below Table 1 and Fig. 2 show significant ROI volumes. The choice of Kruskal-Wallis rather than the ANOVA test is because distributions of ROI volumes are not normal distribution.

**Table 1:** Significant ROI volumes and p-values

| FLDNAME | P-values |
| --- | --- |
| Volume (Cortical Parcellation) of LeftPrecentral | 0.00378 |
| Volume (Cortical Parcellation) of RightLateralOrbitofrontal | 0.004316 |
| Volume (WM Parcellation) of LeftInferiorLateralVentricle | 0.005641 |
| Volume (Cortical Parcellation) of RightPrecentral | 0.008466 |
| Volume (Cortical Parcellation) of LeftParacentral | 0.011357 |
| Volume (Cortical Parcellation) of LeftPrecentral | 0.014085 |
| Volume (WM Parcellation) of LeftVessel | 0.017018 |
| Volume (WM Parcellation) of LeftInferiorLateralVentricle | 0.019348 |
| Volume (Cortical Parcellation) of LeftParacentral | 0.020999 |
| Volume (WM Parcellation) of RightInferiorLateralVentricle | 0.025208 |
| Volume (Cortical Parcellation) of LeftFrontalPole | 0.026786 |
| Volume (Cortical Parcellation) of RightPrecentral | 0.030398 |
| Volume (WM Parcellation) of LeftLateralVentricle | 0.031343 |
| Volume (Cortical Parcellation) of LeftPostcentral | 0.033126 |
| Volume (Cortical Parcellation) of RightTemporalPole | 0.033798 |
| Volume (WM Parcellation) of LeftLateralVentricle | 0.035684 |
| Volume (Cortical Parcellation) of RightTransverseTemporal | 0.048881 |

**Figure 2.**
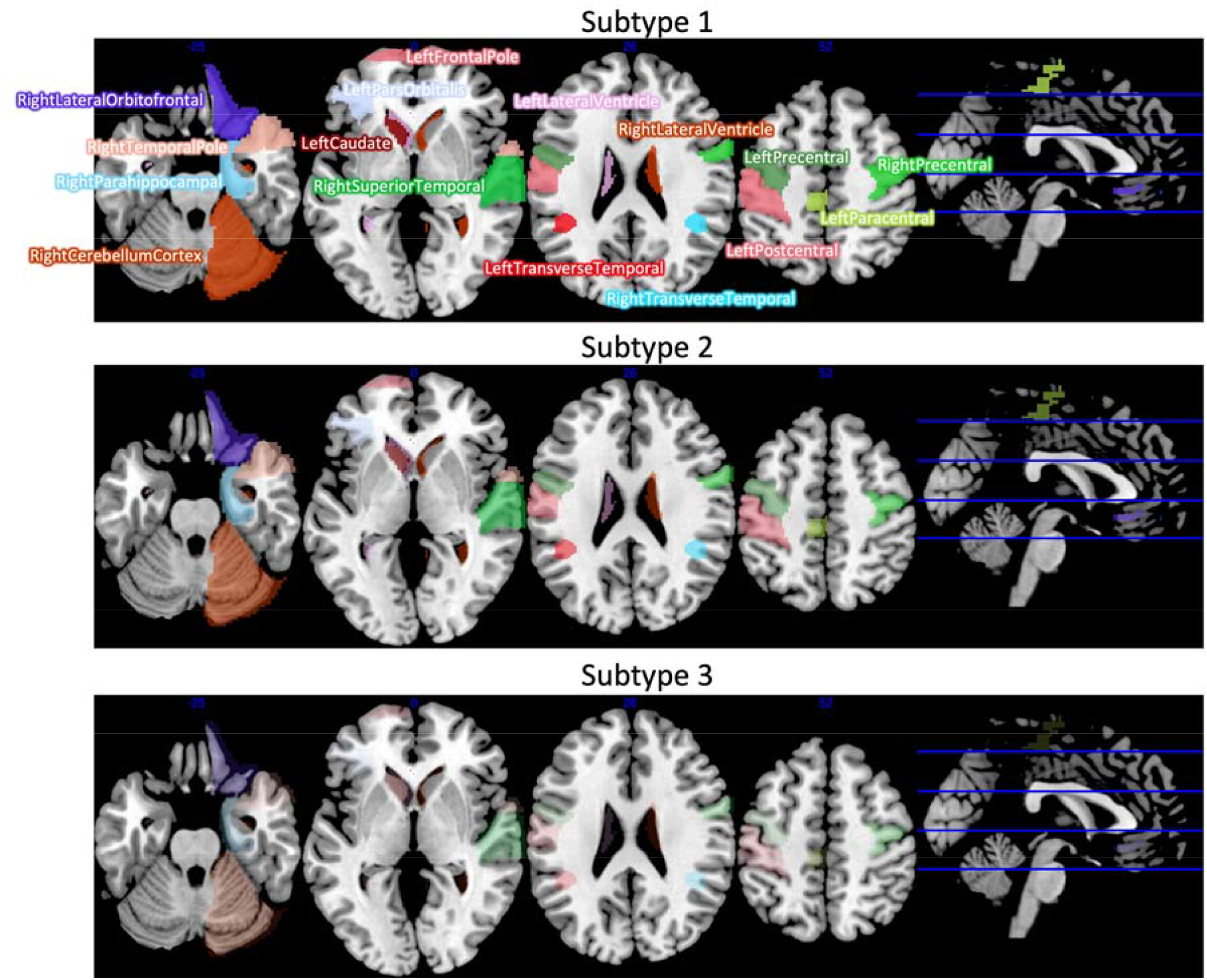
Significant ROI biomarkers

### 2.5 Find significant proteins from subtyping

We use ADNI’s Biomarkers Consortium CSF Proteomics MRM dataset (CSFMRM) to find significantly different proteins between each subtype and normal control (CN) group. The intersected number of participants between 3 subtypes and the CSFMRM are subtype 1 has 10, subtype 2 has 13, and subtype 3 has 20 respectively. While the number of CN participants between the TADPOLE dataset and CSF proteomics is 86.

We first build a contrast matrix between each pair of subtype and CN conditioned on age, gender, and year of education factors to eliminate the bias introduced by these factors. Then we fit a linear model for each pair of subtype and CN and find significant proteins by thresholding |logFC|> 0.2 and Padj < 0.05 from the empirical Bayes’s statistical tests. The p-value (Padj) is corrected by FDR. The actual implementation is via limma library in R. The resulted significant proteins are in Table 2.

**Table 2:** Significant proteins between subtypes and CN.

| Subtype 1 | Subtype 2 | Subtype 3 |
| --- | --- | --- |
| NPTXR | NPTXR | FABPH |
| NPTX2 | FABPH | NPTXR |
| VGF | PIMT | PIMT |
| CA2D1 | KLK11 | GFAP |
| AMD | SCG3 | PCSK1 |
| PTPRN | CH3L1 |  |
| NPTX1 | GFAP |  |
| NRX1A | NPTX1 |  |
| NRX3A | MUC18 |  |
| NRX2A |  |  |
| CCKN |  |  |
| NELL2 |  |  |
| CAD13 |  |  |
| SCG2 |  |  |
| NRCAM |  |  |
| PDYN |  |  |
| PCSK1 |  |  |
| NEGR1 |  |  |
| NCAN |  |  |
| FAM3C |  |  |
| PLDX1 |  |  |
| SE6L1 |  |  |
| CD59 |  |  |
| CSTN1 |  |  |
| CNTN1 |  |  |
| PIMT |  |  |
| CMGA |  |  |
| FABPH |  |  |
| SCG3 |  |  |
| MUC18 |  |  |
| TNR21 |  |  |
| NEO1 |  |  |
| L1CAM |  |  |
| B3GN1 |  |  |
| BTD |  |  |
| NBL1 |  |  |
| SPRL1 |  |  |
| NCAM1 |  |  |
| PVRL1 |  |  |
| CADM3 |  |  |

### 2.6 Find significant pathways from proteins

After we obtain the significant proteins between each pair of subtype and CN, we adopt g:profiler^x^ to perform pathway enrichment analysis and determine statistically enriched gene pathways. We correct p-value of 0.05 threshold with the Benjamini-Hochberg FDR. The gene data sources are GO molecular function, GO cellular component, GO biological process, KEGG, and Reactome. Then we filter output significant pathways by limiting term size between 5 and 350. The resulted pathways between each subtype and CN are listed in Fig 3.

**Figure 3.**
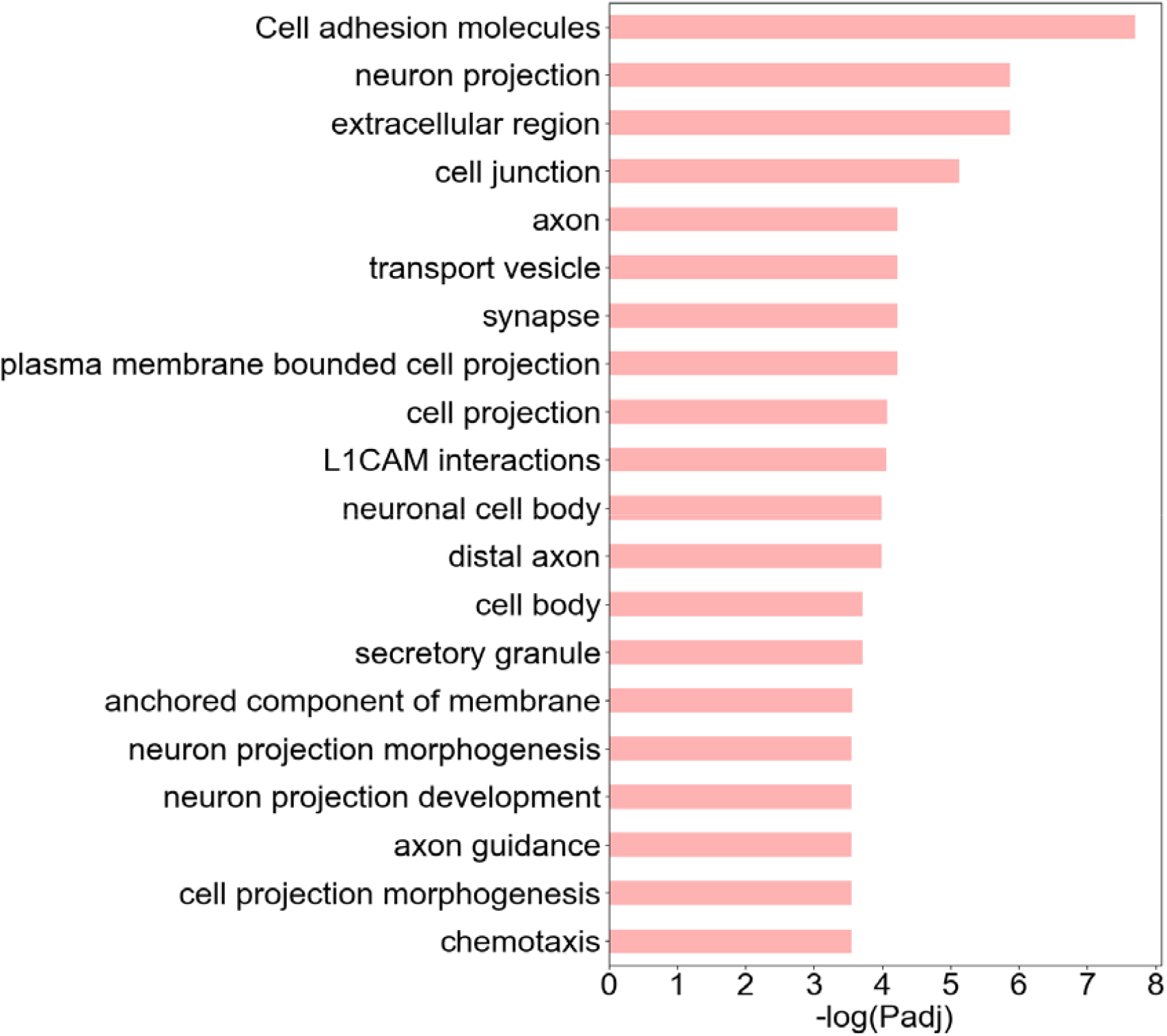
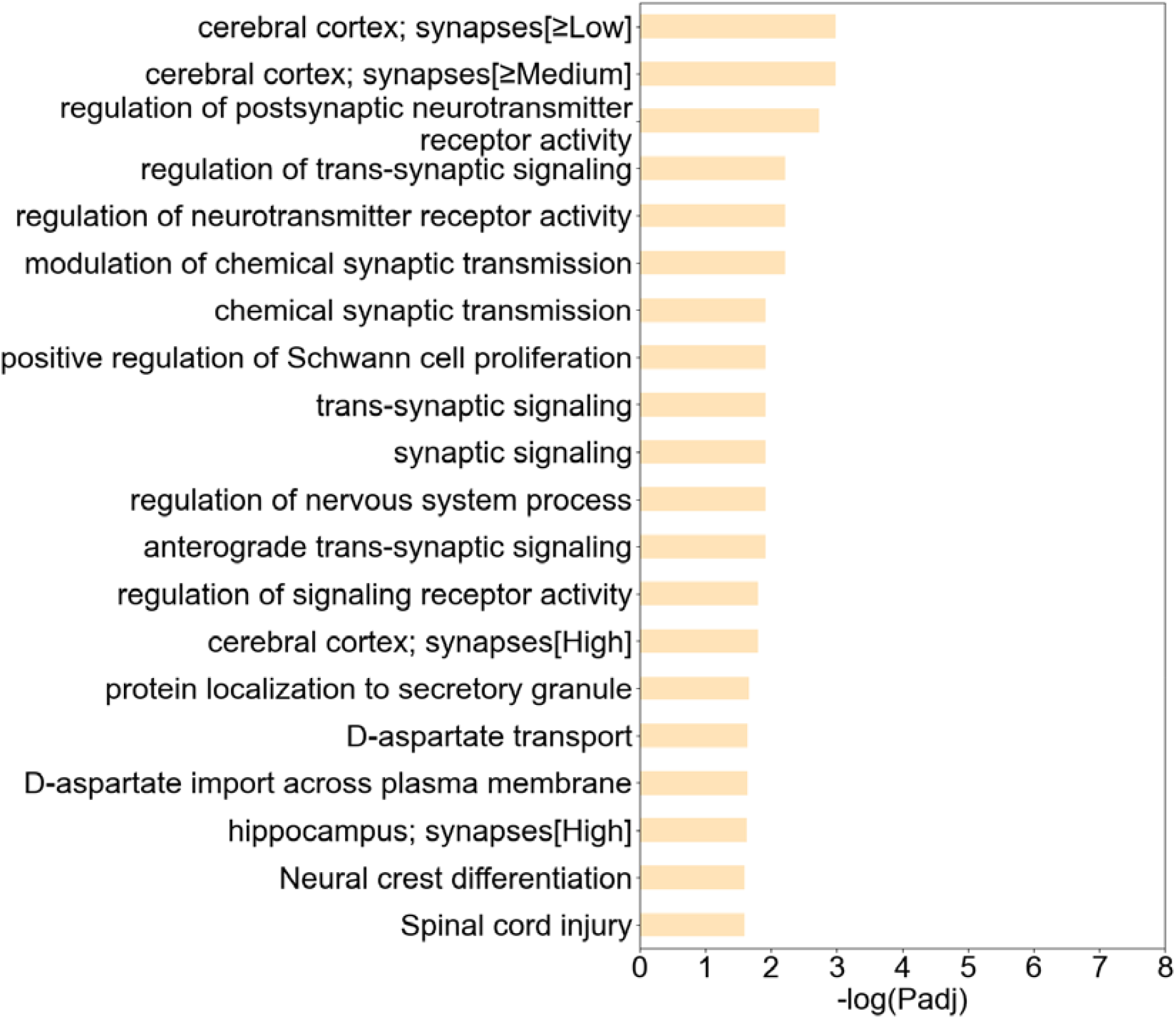
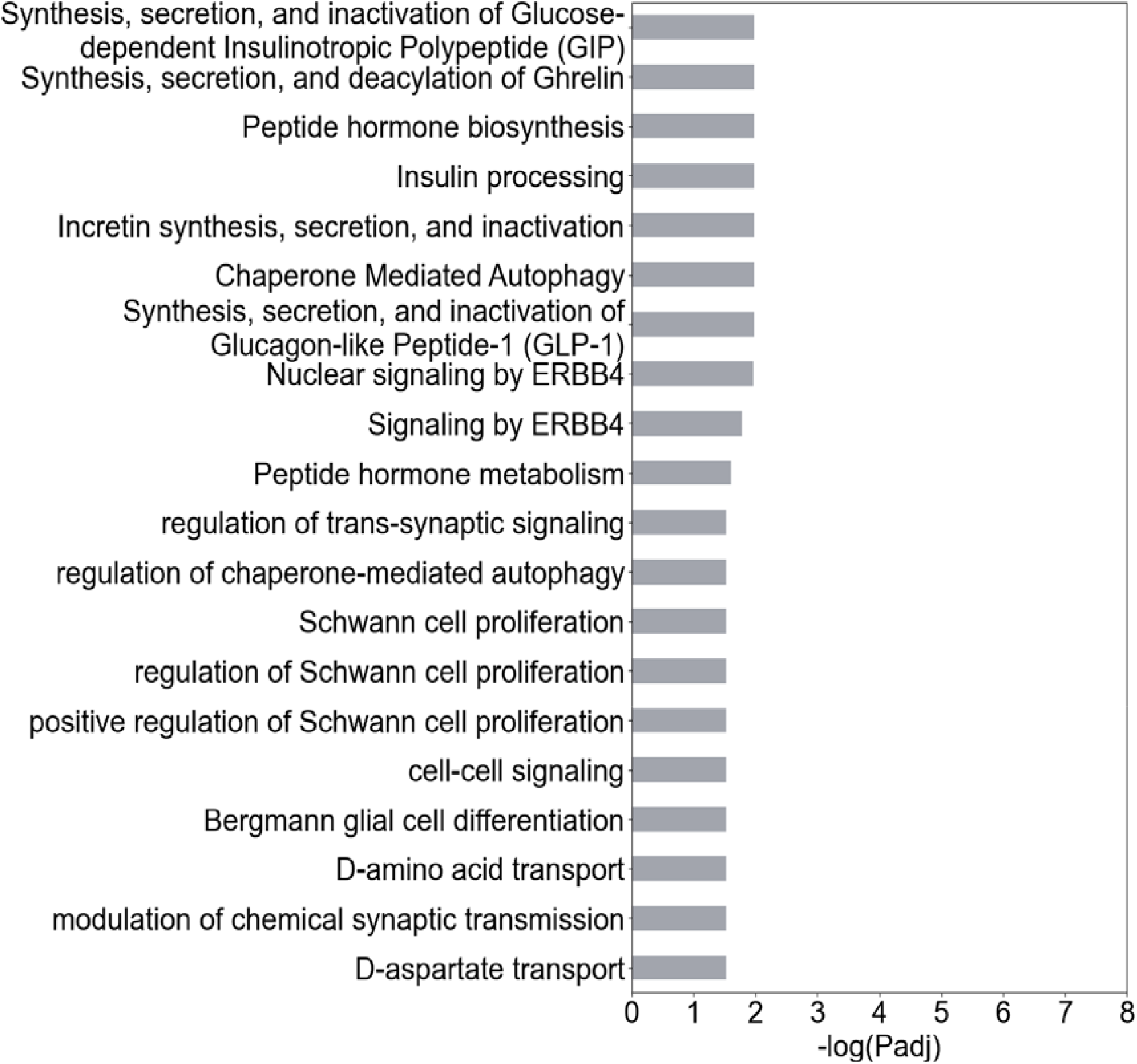
Genetic pathways in 3 subtypes. Red: subtype 1, yellow: subtype 2, gray: subtype 3.

## Data Availability

The ADNI dataset used in this study can be accessed at https://adni.loni.usc.edu/.

## References

i Satone et al., “Learning the Progression and Clinical Subtypes of Alzheimer’s Disease from Longitudinal Clinical Data.”

ii Ferreira et al., “Distinct Subtypes of Alzheimer’s Disease Based on Patterns of Brain Atrophy: Longitudinal Trajectories and Clinical Applications.”

iii Goyal et al., “Characterizing Heterogeneity in the Progression of Alzheimer’s Disease Using Longitudinal Clinical and Neuroimaging Biomarkers”; Su et al., “Comprehensively Modeling Heterogeneous Symptom Progression for Parkinson’s Disease Subtyping.”

iv Faghri et al., “Identification and Prediction of Parkinson’s Disease Subtypes and Progression Using Machine Learning in Two Cohorts.”

v Satone et al., “Predicting Alzheimer’s Disease Progression Trajectory and Clinical Subtypes Using Machine Learning”; Brendel et al., “Comprehensive Subtyping of Parkinson’s Disease Patients with Similarity Fusion: A Case Study with BioFIND Data.”

vi Budgeon et al., “Constructing Longitudinal Disease Progression Curves Using Sparse, Short-Term Individual Data with an Application to Alzheimer’s Disease” Faghri et al., “Identification and Prediction of Parkinson’s Disease Subtypes and Progression Using Machine Learning in Two Cohorts.”

vii Tijms et al., “Pathophysiological Subtypes of Alzheimer’s Disease Based on Cerebrospinal Fluid Proteomics.”

viii Marinescu et al., “Tadpole Challenge: Prediction of Longitudinal Evolution in Alzheimer’s Disease.”

ix Pedregosa et al., “Scikit-Learn: Machine Learning in Python.”

x Reimand et al., “Pathway Enrichment Analysis and Visualization of Omics Data Using g: Profiler, GSEA, Cytoscape and EnrichmentMap.”

